# Prenatal care and clinical characteristics of adolescent pregnancies

**DOI:** 10.64898/2026.11.12.26367235

**Authors:** Angela Victoria Meléndez Loredo, Ricardo Castrejón Salgado, Lisbeth Ruiz Calva

**Affiliations:** Family Medicine, Hospital General Regional con Medicina Familiar No. 1, Instituto Mexicano del Seguro Social, Cuernavaca, Morelos, Mexico; Coordinación de Planeación y Enlace Institucional, OOAD Morelos, Instituto Mexicano del Seguro Social, Morelos, Mexico; Gynecology and Obstetrics, Hospital General Regional con Medicina Familiar No. 1, Instituto Mexicano del Seguro Social, Cuernavaca, Morelos, Mexico

**Keywords:** adolescent pregnancy, prenatal care, reproductive health, medical records, family medicine

## Abstract

**Background:** Descriptions of adolescent pregnancy can identify priorities for prenatal care and postpartum follow-up. This study characterized prenatal care and clinical outcomes documented in adolescent pregnancy records.

**Methods:** We conducted a retrospective, descriptive, observational cross-sectional review of records covering 2020–2024 at a single family medicine outpatient service. All eligible records were included. Prenatal care and clinical characteristics were summarized using frequencies and percentages. Reporting was restricted to aggregate results to reduce the risk of participant identification.

**Results:** Thirty-nine records were analyzed. The first prenatal visit occurred in the second trimester in 53.85%. Overweight or obesity was recorded in 71.8%. Urinary tract infection occurred in 66.67% and gestational hypertension in 28.21%. No preterm births or obstetric hemorrhages were recorded. Vaginal delivery accounted for 51.28% and cesarean delivery for 48.72%. Postpartum contraceptive method was unspecified in 64.10% of records.

**Conclusions:** This small, single-center study identified frequent urinary tract infections, a high recorded proportion of overweight or obesity, prenatal entry commonly after the first trimester, and incomplete postpartum contraceptive documentation. These findings identify local priorities for care and record quality; they do not establish causal relationships or population prevalence.

## Introduction

Adolescent pregnancy has health, educational and social implications. The World Health Organization identifies barriers to contraception and unequal access to maternal care as important concerns for adolescents, alongside increased risks of several maternal and neonatal complications [1]. These concerns support the need for local descriptions of the adolescents who reach health services and of the care documented in their records.

Prenatal service use varies across adolescent populations. A Brazilian study found differences in prenatal attendance between younger and older adolescents [2]. Findings from other settings cannot be assumed to describe the population attending an individual family medicine service, supporting the value of carefully reported local evidence.

Describing prenatal entry and clinical outcomes recorded for adolescents may help identify priorities for service improvement. The objective of this report was to characterize prenatal care and clinical outcomes documented in adolescent pregnancy records during 2020–2024, while limiting disclosure of participant characteristics.

## Methods

### Study design and setting

This was a retrospective, descriptive, observational cross-sectional study based on medical record review at a single family medicine outpatient service. Records concerned pregnancies during 2020–2024. Data extraction was performed after institutional committee approval. The service location and detailed extraction dates are not reported to reduce the risk of participant identification.

### Eligibility and record selection

Records were eligible if they concerned adolescents aged 10–19 years who experienced pregnancy during the study period and were registered in the family medicine outpatient service. Incomplete records or records with insufficient information on pregnancy or variables of interest were excluded. All records meeting the selection criteria were included, without a sample size calculation or sampling procedure. The final study population comprised 39 records. The total number screened and the number excluded were unavailable for reporting.

### Data sources and variables

The research team obtained the adolescent pregnancy census from the outpatient service and reviewed records in the Sistema de Información de Medicina Familiar (SIMF). A structured collection form was used, and information was entered into Microsoft Excel. Baseline information was extracted from the first prenatal note. Subsequent clinical notes and paper records were reviewed to ascertain pregnancy complications, delivery outcomes and postpartum contraception.

The results presented here concern trimester at the first prenatal visit, nutritional status at the beginning of pregnancy, elevated blood pressure at the first visit, gestational hypertension, urinary tract infection, preterm birth, obstetric hemorrhage, mode of delivery and postpartum contraceptive method. Other demographic and reproductive history variables collected for the original study are not presented in this public report for confidentiality.

Nutritional status at the beginning of pregnancy was classified as normal weight, overweight or obesity according to body mass index (BMI), calculated as weight in kilograms divided by height in meters squared using measurements recorded in SIMF. The original classifications were retained; the specific BMI cutoffs and age reference were not available for reporting. Preterm birth was defined as birth before 37 completed gestational weeks. Clinical conditions were abstracted from documented diagnoses; no independent diagnostic adjudication was reported. Trimester at first prenatal contact was estimated retrospectively from recorded dates.

### Statistical analysis

The database was cleaned and organized in Microsoft Excel and analyzed in Stata version 14.0. Categorical variables are summarized as counts and percentages. The denominator is 39 throughout; an unspecified postpartum contraceptive method is retained as a separate category. No bivariate or multivariable analyses were conducted. Published counts and percentages for the retained clinical variables are unchanged. Exact participant ages, age summary statistics, ages at reproductive events, occupation, education, marital status, gravidity, number of sexual partners and pre-existing medical history are omitted from this report. Nonzero outcome categories with fewer than five observations are not disclosed, and complementary categories that would permit their reconstruction are not presented. Tables contain only separate marginal distributions, without participant-level records or cross-tabulations linking characteristics.

### Ethics

The protocol was approved on 22 May 2025 by the Comité Local de Investigación en Salud No. 1701 at Hospital General Regional con Medicina Familiar No. 1, IMSS, under institutional registration R-2025-1701-064, with review by the institutional Comité de Ética en Investigación. The ethics committee approved a waiver of informed consent for this retrospective medical record review. The study involved no intervention. Names, social security numbers and other direct identifiers were not included in the data collection form, and confidentiality was maintained.

## Results

The analysis included 39 records. The first prenatal visit occurred in the first trimester in 12 cases (30.77%), second trimester in 21 (53.85%), and third trimester in six (15.38%) (Table 1).

**Table 1.** Timing of the first prenatal visit. N = 39. Prenatal trimester was estimated retrospectively from recorded dates. Counts are aggregate totals and are not linked to individual characteristics.

| Characteristic | n | % |
|---|---|---|
| First | 12 | 30.77 |
| Second | 21 | 53.85 |
| Third | 6 | 15.38 |

Normal weight was recorded in 11 adolescents (28.21%), overweight in 17 (43.59%) and obesity in 11 (28.21%). Five (12.82%) had elevated blood pressure at the first visit. Gestational hypertension was recorded in 11 (28.21%) and urinary tract infection in 26 (66.67%). No preterm births or obstetric hemorrhages were recorded (Table 2).

**Table 2.**
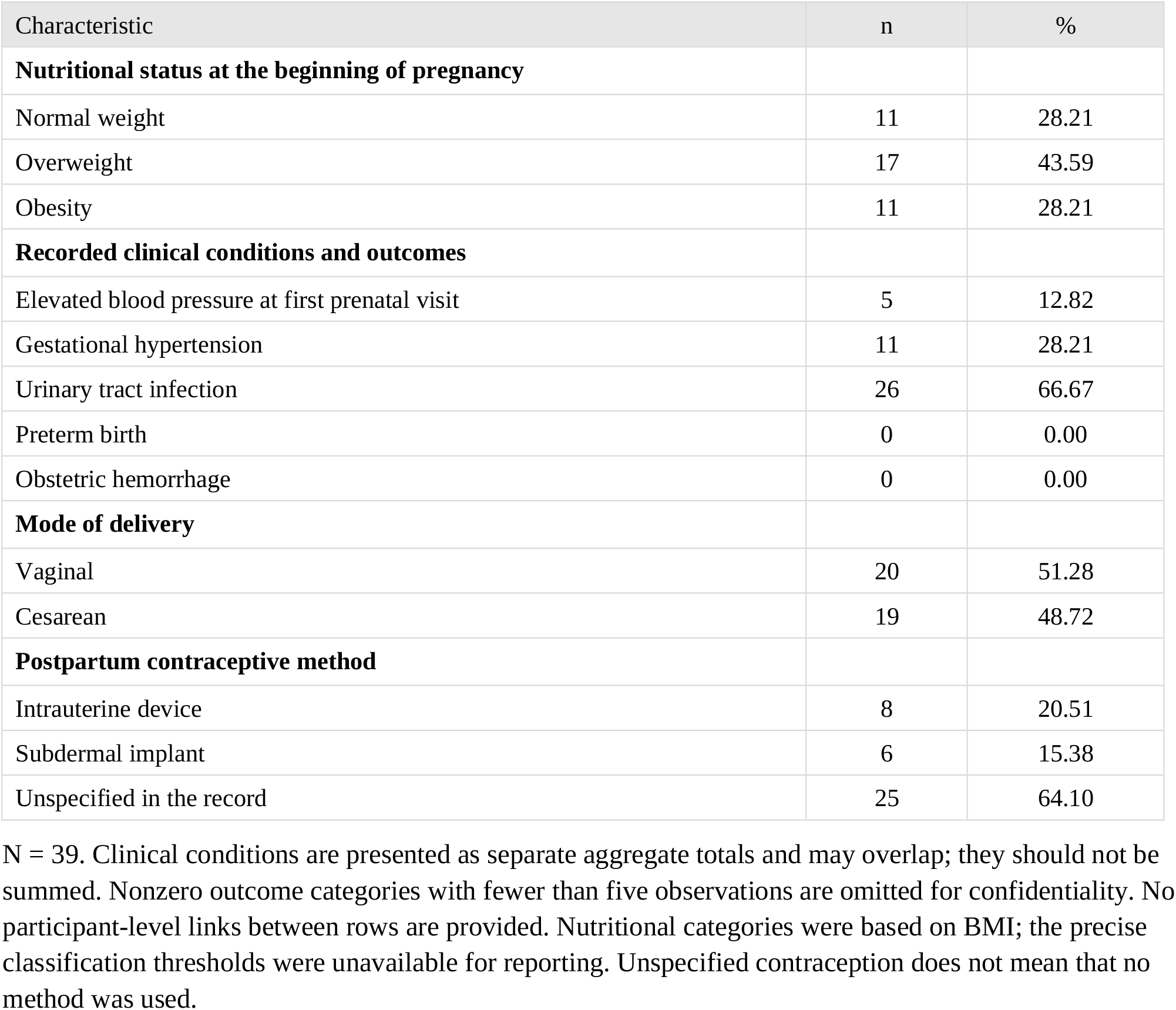
Clinical characteristics and obstetric outcomes.

Twenty adolescents (51.28%) had vaginal deliveries and 19 (48.72%) had cesarean deliveries. Postpartum contraception was documented as an intrauterine device in eight cases (20.51%) and a subdermal implant in six (15.38%). The method was unspecified in 25 records (64.10%) (Table 2).

## Discussion

This review describes prenatal care and clinical characteristics in an adolescent population. The main findings were frequent recorded urinary tract infections, a high proportion classified as overweight or obese, prenatal entry most commonly in the second trimester, and substantial missing documentation of postpartum contraceptive method. These observations describe records available at one service and should not be interpreted as population estimates.

Most first prenatal contacts occurred after the first trimester. Assis and colleagues found fewer than six prenatal visits more frequently among younger than older adolescents in Brazil [2]. That measure differs from trimester at first contact, but both findings draw attention to prenatal service use. The present study did not measure barriers to attendance, care received elsewhere, or the number and content of subsequent visits. Consequently, later first contact at this unit cannot establish the cause of delayed entry or the adequacy of the entire prenatal course.

Urinary tract infection was the most frequently documented clinical condition. Its frequency should be interpreted in light of the retrospective design and the absence of a reported standardized case definition or microbiological confirmation procedure. Likewise, the coexistence of overweight or obesity and hypertensive disorders does not demonstrate an association between them. Nutritional classifications require particular caution because the available information does not specify how adolescent age or gestational timing was incorporated into the body mass index interpretation.

Cesarean delivery accounted for 48.72% of cases. A systematic review in the Eastern Mediterranean region reported a pooled cesarean delivery proportion of 15.9% [3]. Differences in settings, selection and definitions preclude interpreting that figure as a benchmark for this service. The present data do not include cesarean indications and cannot establish whether individual procedures were appropriate. The absence of recorded preterm birth and hemorrhage in 39 records does not rule out these outcomes in the wider service population.

Postpartum contraceptive method was unspecified in almost two thirds of records. This is a documentation finding, not evidence that these adolescents received no contraception. Clear recording of counseling, method provision and follow-up would allow a more reliable assessment of postpartum reproductive care. Future work should distinguish nonuse, refusal, provision elsewhere and missing documentation.

### Strengths and limitations

The study brings together prenatal care and clinical information from a family medicine population and includes all records meeting the stated criteria. However, the small, single-center population limits precision and generalizability. Exclusion of incomplete records may introduce selection bias, and the numbers screened and excluded were unavailable. Routine records were not designed for research, and retrospective estimation of prenatal trimester may cause misclassification. Standardized diagnostic verification and the precise BMI classification thresholds were unavailable for reporting. No comparison group or association analyses were available, and causal conclusions are not supported. Confidentiality-related omissions limit the description of demographic variation, reproductive history and uncommon outcomes. Omitted information should not be interpreted as absence of the corresponding characteristics or events.

## Conclusions

Among 39 adolescent pregnancy records reviewed for 2020–2024, prenatal entry was commonly documented in the second trimester. Urinary tract infection and recorded overweight or obesity were frequent. The findings support review of prenatal entry, clinical documentation and postpartum contraceptive recording. Larger studies with explicit selection flow and standardized definitions are needed to assess the broader population and relationships between clinical characteristics and outcomes.

## Data Availability

All data produced in the present study are available upon reasonable request to the authors

## Declarations

### Ethical approval and consent

The study was approved under institutional registration R-2025-1701-064. The ethics committee approved a waiver of informed consent for the retrospective record review. No individual identifying information is presented.

### Funding

This research received no external funding.

### Competing interests

The authors declare no competing interests.

### Data availability

Aggregate data suitable for public disclosure are presented in the manuscript tables. Requests for additional de-identified data may be directed to the corresponding author and will be considered under institutional data protection requirements and a participant confidentiality assessment. Information withheld from this public report for confidentiality is not supplied in supplementary files.

### Author approval

All authors have read and approved the manuscript and agree to its submission to medRxiv.

### Clinical trial registration

Not applicable. This was an observational medical record review without an assigned intervention. The SIRELCIS number is an institutional research registration, not a clinical trial registration.

### Use of artificial intelligence

ChatGPT was used to assist with English translation, manuscript drafting and formatting based on the source thesis.

## Notes

### Competing Interest Statement

The authors have declared no competing interest.

### Author Declarations

The protocol was approved on 22 May 2025 by the Comité Local de Investigación en Salud No. 1701 at Hospital General Regional con Medicina Familiar No. 1, IMSS, under institutional registration R-2025-1701-064, with review by the institutional Comité de Ética en Investigación.

